# Reconstruction of immunisation during conflict: A mixed-methods cohort evaluation of programme delivery and outcomes in Myanmar

**DOI:** 10.64898/2026.05.15.26352743

**Authors:** Daniel Fishbein, Hein Thura Aung, Roxy Ong, Aurora Nyein, Zarni Lynn Kyaw, Emily Karenni, Jie Jie, Kanyar Maw, Kaung Khant, April Poe, MayThandi Win, Brianna Grissom, Cynthie Tinoo

## Abstract

**Introduction:** Routine childhood immunisation is frequently disrupted in conflict-affected settings, leaving many children unvaccinated (zero-dose [ZD]). Their vaccination is now a global priority, but published evidence on restoring immunisation services in these settings is limited. We evaluated a nurse-led, community-based Expanded Programme on Immunisation adapted to a conflict-affected setting in Myanmar, focusing on factors associated with full immunisation (FI) among ZD children.

**Methods:** This mixed-methods observational cohort study enrolled children from November 2023 to December 2025; analyses of FI outcomes were restricted to children enrolled ≥18 months, with primary analyses focused on ZD children. Associations between programme delivery factors—including vaccination opportunity (the ratio of vaccination sessions available to visits required for FI based on age and vaccination schedule [accelerated versus routine])—and FI were assessed using mixed-effects logistic regression with a random intercept for site. Programme cost and qualitative data from document review and questionnaires were also analysed.

**Results:** Of 13,263 children enrolled, 6563 (49%) were in the analytic cohort; 2,684 (20%) were ZD. Among ZD, 452 (17%) were FI at 12 months and 1329 (50%) at 18 months. Accelerated schedule (OR 3.00, 95% CI 1.11–8.13) and greater vaccination opportunity (OR 2.1 per 0.5 unit increase in opportunity, 95% CI 1.8–2.4) were strongly associated with FI at 12 months, with smaller effects at 18 months. The cost per fully immunised ZD child was US$147, primarily reflecting substantial vaccine costs. Qualitative findings indicate that community engagement increased demand and access, but insecurity and logistical challenges limited service continuity and vaccination opportunities.

**Conclusion:** FI improved over time but remained suboptimal through 18 months. Vaccination opportunity and schedule influenced the timing of FI, but sustained follow-up was critical for completion. Community-based delivery enabled restoration of immunisation services where formal systems had collapsed, demonstrating what is possible—and what it demands—in active conflict.

**Funding:** United Nations

**KEY MESSAGES:** *WHAT IS ALREADY KNOWN ON THIS TOPIC:* Reaching zero-dose (ZD) children in fragile and conflict-affected settings is a priority of the WHO Immunization Agenda 2030, but published evidence on programmes attempting to restore immunisation in such settings is limited. Routine immunisation services in Myanmar collapsed following the 2021 military coup and subsequent access restrictions amounted to a de facto humanitarian blockade.

*WHAT THIS STUDY ADDS:* This study provides among the first longitudinal evidence on implementation, costs, and outcomes of community-based immunisation among ZD children during active conflict. Half of ZD children and 78% of those who were incompletely immunised at enrolment achieved full immunisation within 18 months. Programme factors including accelerated vaccination schedule and increased vaccination opportunities were associated with earlier completion; sustained retention was the dominant determinant of overall coverage.

*HOW THIS STUDY MIGHT AFFECT RESEARCH, PRACTICE OR POLICY:* Community-based delivery through civil society organisations can restore immunisation services in high priority settings but its fragility and need for sustained external support cannot be ignored. Expanding vaccination opportunities, using accelerated schedules, and actively retaining children after enrolment are actionable steps to increase coverage.

## BACKGROUND

Childhood vaccination through the Expanded Programme on Immunisation (EPI) remains one of the most effective public health interventions, yet global progress has stalled.^1-4^ The World Health Organization’s Immunization Agenda 2030 (IA2030) identifies zero-dose (ZD) children—operationally defined as those who have not received the first dose of a diphtheria containing vaccine—as a key marker of inequity.^2^ These children are concentrated in conflict-affected and fragile settings, where overlapping socioeconomic, geographic, and health-system barriers are compounded by disrupted, politicised, or inaccessible routine services.^4-7^

Rel71establishing routine immunisation in such areas is widely recognised as a major challenge in global health.^4 8^ Recognising this complexity, Gavi, the Vaccine Alliance, developed the Zero-Dose Immunisation Programme (ZIP) and a Zero-dose Learning Centre^9^ to assist in strengthening humanitarian partnerships and expanding immunisation delivery in fragile settings.^10 11^ These initiatives now form part of Gavi’s broader Gavi Leap strategy to accelerate progress toward IA2030 equity goals in fragile and conflict-affected settings.^11^

Despite these efforts, IA2030 targets—including halving the global number of ZD children—are unlikely to be met without major shifts in service delivery in settings with the greatest inequity.^2 4 12^ Although ZD populations are increasingly prioritised, most studies rely on cross-sectional coverage estimates and risk factors, rather than longitudinal assessments of schedule completion, and few evaluate delivery in active conflict settings.^13-15^ This represents a critical evidence gap.

Myanmar provides a relevant case study. Long-standing instability contributed to low immunisation coverage, which worsened following the 2021 military coup, as the health system fragmented and humanitarian access declined.^16-18^ Unwilling to work for the military government, many health workers joined the Civil Disobedience Movement and relocated to ethnic minority–controlled areas.^19 20^ Although efforts to restore routine immunisation in these areas were made, information is limited by fear of disclosure and lack of vaccines and other resources.^19 20^

In 2023, a nurse-led pilot programme in Karenni State demonstrated that immunisation could be re-established using cross-border vaccine procurement and community-based delivery.^20^ This pilot informed the present evaluation within a larger programme targeting underserved children in an active conflict setting.^21 22^ This longitudinal cohort study evaluated determinants of full immunisation (FI), including baseline characteristics, programme exposure, and retention, with a prespecified target of 70% FI at 12 months after enrolment.

## METHODS

The evaluation was conducted by investigators involved in programme implementation; primary outcomes and analytic objectives were prespecified, while details of the modelling approach were finalised during analysis.

### Study design and setting

The complexity of analysing this attempt to restore routine childhood immunisation in conflict-affected settings necessitated a comprehensive research approach that captures both measurable outcomes and contextual factors. Using a mixed-methods design allows this study to integrate quantitative and qualitative data, providing a nuanced understanding of programme delivery and immunisation outcomes.

The quantitative component was an observational cohort study of programme implementation and immunisation outcomes during a nurse-led vaccination programme in central and western Karenni State, Myanmar, a conflict-affected region with disrupted state health services following the military coup.^23 24^ The immunisation programme was delivered by the Karenni Nurses Association (KNA), a civil society organisation of Civil Disobedience Movement^20^ health professionals providing primary care across five resistance-controlled townships (Supplementary Figure 1). Age-specific denominators were unavailable due to lack of a reliable census since 2014.

Vaccines were procured at commercial prices in Thailand (Table 1. Comparison of vaccine schedules and antigen prices) and transported via cross-border routes (Supplementary Figure 2). Eighteen vaccination sites were established. Sessions were held monthly where feasible, depending on vaccine availability, road conditions, and safety (Supplementary Figure 3).

**Table 1.** Comparison of vaccine schedules and antigen prices.

| Vaccine | Accelerated/catch-up schedule, programme | Routine schedule, programme | Routine schedule, Myanmar, 2024 | Antigen price, Thailand (US\$) | Antigen price, Gavi (US\$)<sup>31</sup> |
| --- | --- | --- | --- | --- | --- |
| BCG | Birth–2 months (up to 1 year) | Birth–2 months (up to 1 year) | Birth–2 months | 1.27 | 0.20 |
| Hepatitis B (birth dose) | Not included* | Not included* | Birth | Not available | 0.24 |
| Pentavalent | 3 monthly doses from age ≥2 months | 2, 4, and 6 months | 2, 4, 6, and 18 months | 13.73 | 0.84 |
| Oral polio | 3 monthly doses from age ≥2 months | 2, 4, and 6 months | 2, 4, and 6 months | 1.67 | 0.14 |
| Injectable polio | Not included<sup>†</sup> | Not included<sup>†</sup> | 4 months | Not available | 2.10 |
| Measles, mumps, rubella | 2 monthly doses from age ≥9 months | 9 and 18 months | 9 and 18 months<sup>§</sup> | 13.44 | 0.82<sup>‡</sup> |
| Pneumococcal conjugate | Not included* | Not included* | 6, 10, and 14 weeks | 3.50 | 2.75 |
| Japanese encephalitis (live attenuated) | 1 dose from age ≥9 months<sup>¶</sup> | 9 months<sup>¶</sup> | 9 months | 10.59 | 0.45 |
| Rotavirus | Not included* | Not included* | 2 and 4 months | Not available | 2.22 |
| Td (prenatal) | First contact and 4 weeks later | First contact and 4 weeks later | First contact and 4 weeks later | 0.47 | 0.25 |
| Vitamin A | Not included* | Not included* | Twice per year | Not applicable | Not included |
| HPV | Not included* | Not included* | 9-year-old girls | Not available | 4.50 |

\* Included in the Myanmar EPI schedule but not included in the programme because funding not provided
† Became available in 2025 but not evaluated because of insufficient follow up before end of evaluation
‡ Gavi prices are for measles rubella vaccine
§ Myanmar national schedule is for measles rubella vaccine only
¶ Discontinued by United Nations in January 2025

To minimise bias, outcomes were pre-specified, active outreach was used to identify and encourage enrolment of eligible children, standardised definitions and data collection procedures were applied across sites, and mixed-effects models were used to account for clustering and contextual variation.

### Participants and eligibility

All children aged <5 years presenting for vaccination were included. Data routinely collected in Myanmar’s EPI—including demographic information—name, date of birth, age at enrolment, sex, residence (internally displaced persons [IDP] camp or village)— and date of enrolment (defined as the initial vaccination visit) and vaccines received—were recorded in register books and entered into a Burmese-language Microsoft Access database with automated quality checks. Age at enrolment was calculated from date of birth to the first vaccination administered by the programme.

Children were classified at enrolment as ZD if they had not received a first dose of pentavalent vaccine, and as incompletely immunised if they had received at least one dose. Consistent with recent definitions in IA2030 documents,^8^ WHO under-immunisation definitions were not used, as children receiving pentavalent dose 3 could be considered fully immunised under WHO criteria^25^ but not under this study’s definition, which required complete doses of all five schedule vaccines.

Active prospective follow-up was not possible given security issues and difficulty and risk of locating children in IDP camps and communities. Rather, outcomes were ascertained retrospectively using vaccination records of children enrolled ≥18 months before the evaluation period ended (analytic cohort) to ensure equal opportunity for follow-up and avoid immortal time bias. Analyses focused on ZD children; outcomes 12- and 18-month after enrolment are reported for this cohort unless otherwise specified.^8^

### Programme delivery and exposure variables

Due to funding constraints, the programme provided only five (Bacillus Calmette–Guérin [BCG], oral polio [OPV], measles, mumps, and rubella [MMR], Japanese encephalitis [JE]) of Myanmar’s nine-vaccine schedule (Table 1. Comparison of vaccine schedules and antigen prices).^26^ The microplan specified the use of Myanmar’s accelerated catch-up schedule, although some sites later switched to Myanmar’s routine schedule. JE vaccine provision was discontinued by the United Nations in January 2025 due to funding shortages.

Variation in vaccination sessions offered across sites reflected differences in conflict, access, and vaccine availability, and was quantified using programme session records (Supplementary Figure 3). For each child, sessions available were defined as the number of vaccination sessions conducted at the child’s enrolment site from the date of enrolment until achievement of FI or 18 months after enrolment, whichever occurred first. A vaccination opportunity index was then calculated as the ratio of sessions available to the number of visits required to complete the vaccination schedule (Supplementary table 1). Standardisation was necessary because the number of visits required varied by age at enrolment and vaccination schedule.

### Outcomes

Standard UN indicators were not well suited to this setting (Supplementary table 2), so programme-specific outcomes were defined. The primary outcome was full immunisation (FI), defined as receipt of all scheduled pentavalent, OPV, MMR, and JE doses meeting minimum age and interval requirements. The KNA team used a modified Delphi process to set a target of 70% FI at 12 months, consistent with the 12-month FI rate observed in the pilot study.^20^ Timing of FI was based on the date of the final required dose within the programme. Sensitivity analysis assessed the impact of JE discontinuation.

Secondary outcomes included FI at 18 months, time to FI, retention, and dropout. Retention at 12 months was defined as return for at least one vaccination visit within one year. Dropout was defined as failure to return within specified time intervals (3, 6, 9, and 12 months). A time-based definition was used to accommodate heterogeneous enrolment ages and vaccination status.

### Data analyses

#### Quantitative

Descriptive analyses summarised demographic characteristics and outcomes. Group comparisons used Fisher’s exact and non-parametric tests (Wilcoxon rank-sum or Kruskal–Wallis). Kaplan–Meier methods described time to FI.

Return visits at 3, 6, 9, and 12 months after enrolment were evaluated as predictors of immunisation status at 18 months (i.e., early indicators of later dropout) using sensitivity, specificity, positive predictive value, and negative predictive value.

Mixed-effects logistic regression models were used to estimate adjusted associations with FI, including a random intercept for site to account for clustering. Fixed effects included demographic characteristics, vaccination schedule (accelerated vs routine), and programme-related measures. We applied a sequential modelling framework: Model A included baseline characteristics (age, sex, residence); Model B additionally incorporated programme exposure using a vaccination opportunity index reflecting variation in service availability and individual opportunity for vaccination. Model C added programme retention at 12 months as a measure of sustained engagement. This approach allowed assessment of the relative contribution of baseline factors, programme exposure, and retention to immunisation outcomes. Results are presented as odds ratios (ORs) with 95% confidence intervals.

#### Qualitative

Qualitative analysis was undertaken to contextualise quantitative findings and identify operational, governance, and community-level factors influencing vaccination service delivery. Programme records from November 2023 to December 2025 were analysed using a document analysis approach,^27 28^ including eight quarterly reports, 29 monthly reports, and one programme success report. Questionnaire responses from vaccination nurses in April 2025, based on a strengths, weaknesses, opportunities, and challenges (SWOT) framework, were included to capture frontline perspectives. Photographs of vaccine transportation were reviewed to support interpretation of logistical constraints.

Codes were grouped into themes reflecting programme coordination and governance, operational and logistical challenges, access barriers, and contextual factors using QDA Miner Lite. Reflexive thematic analysis was applied to identify and interpret recurring patterns related to facilitators and barriers influencing both operational and demand-side aspects of service delivery.^29 30^

Cost analysis. Programme cost data were obtained from financial records and classified into vaccine and delivery costs using UNICEF’s costing framework.^1^ Costs per dose, per fully immunised child, and per enrolled child were calculated, with sensitivity analyses using reference vaccine prices.

Patient and public involvement. Patients and the public were not formally involved. The programme incorporated community engagement, including collaboration with village leaders, camp committees, and volunteers to support identification of children and delivery of services.

### Ethical approval

Formal review through a Myanmar-based institutional ethics committee was not feasible because of conflict-related security constraints. Approval was obtained from the Mae Tao Clinic Community Ethics Advisory Board (CEAB-2025-004). Procedures followed the Declaration of Helsinki. Data were stored in a secure, password-protected database.

Use of artificial intelligence. ChatGPT (OpenAI) was used to support literature searches, R programming, and manuscript editing. All outputs were reviewed and verified by the authors.

## RESULTS

### Quantitative analyses

#### Cohort characteristics

A total of 13,263 children were enrolled. Of these, 6563 (49%) were eligible for the analytic cohort (Figure 1. Participant flow diagram of enrolled children and vaccination outcomes in the analytic cohort). Compared with children in the analytic cohort, those with <18 months elapsed since enrolment were younger and more likely to be ZD (Supplementary table 3). Over time, age at enrolment decreased and the proportion of ZD children increased (Supplementary figure 4). Detailed stratified comparisons, including characteristics of the ZD group that is the focus of subsequent analyses, are shown in Supplementary table 4.

**Figure 1.**
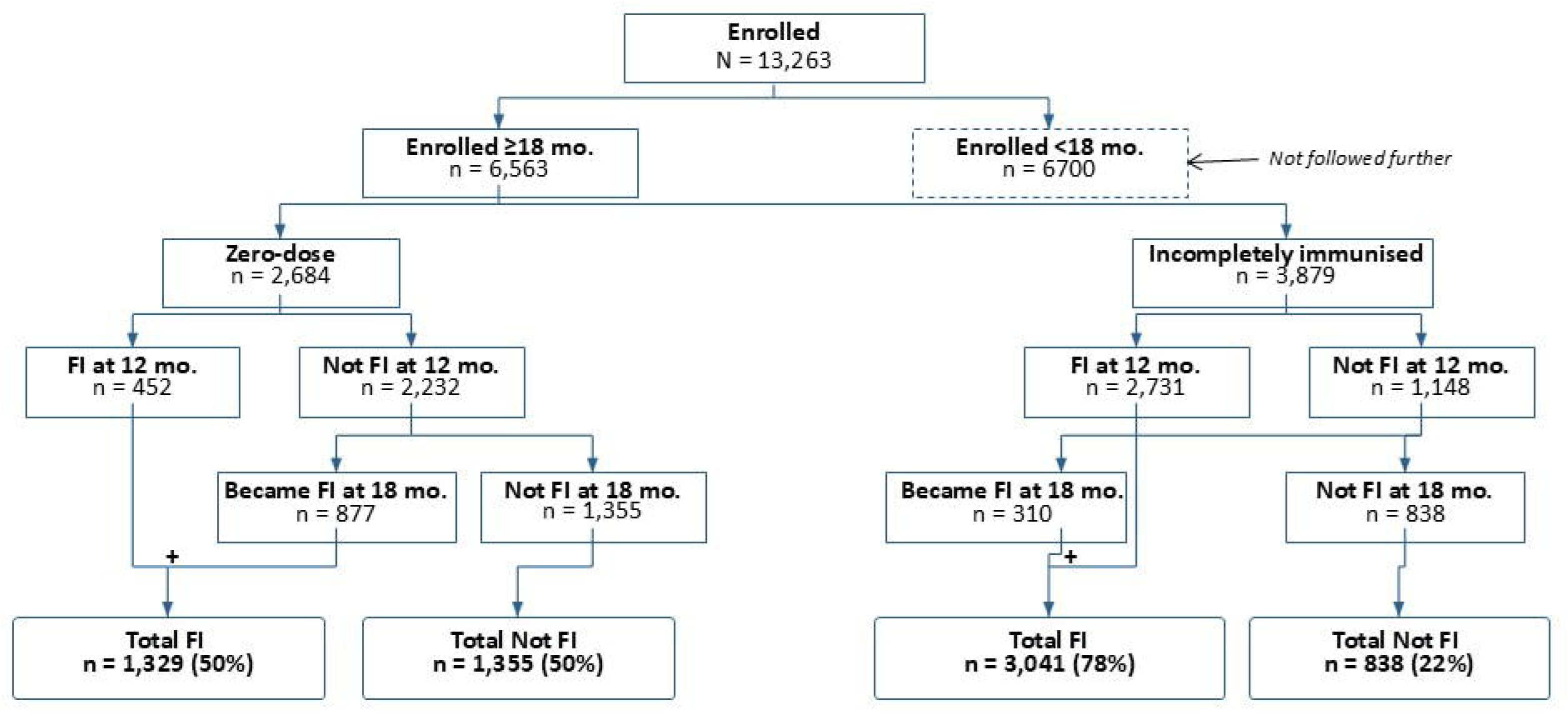
Participant flow diagram of enrolled children and vaccination outcomes in the analytic cohort. Children were classified at enrolment as zero-dose (no prior diphtheria-containing vaccine) or incompletely immunised (prior diphtheria-containing vaccine). Each group was stratified by time since enrolment (<18 months or ≥18 months). Those with ≥18 months since enrolment constituted the analytic cohort. Full immunisation (FI), defined as receipt of all required doses of pentavalent, oral polio, measles–mumps–rubella (MMR), and Japanese encephalitis vaccines at 12 and 18 months after enrolment is shown. Percentages are calculated based on the total number of zero-dose and incompletely immunised children in the analytic cohort.

Univariate analyses of outcomes. Among ZD children, full immunisation increased from 17% at 12 months to 50% at 18 months after enrolment (Figure 1. Participant flow diagram of enrolled children and vaccination outcomes in the analytic cohort). Compared with ZD children, FI was higher among incompletely immunised children at both 12 months (70%) and 18 months (78%). Time to FI also differed substantially by baseline status (log-rank p<0.001). Among incompletely immunised children, FI was achieved rapidly (median 32 days, 95% CI 31–33), whereas the median was not reached among ZD children within 18 months. Differences between groups were most pronounced early and narrowed over time (Supplementary Figure 5). In a sensitivity analysis, exclusion of JE vaccine from FI increased FI by 3% at 12 months and 8% at 18 months.

Several characteristics differed between ZD children who did and did not achieve FI at 12 months and 18 months after enrolment (table 2. Factors associated with full immunisation among ZD children at 12 and 18 months after enrolment). Children who achieved FI at 12 months were older at enrolment (median 21 vs 3 months, p<0.001), more likely to have been vaccinated at sites using accelerated schedules (30% vs 20%, p<0.001), and slightly less likely to reside in IDP settings (37% vs 45%, p=0.005). At 18 months, associations with age and exposure to accelerated schedules persisted, albeit somewhat less strongly, while residence was no longer associated with FI.

**Table 2.** Factors associated with full immunisation among zero-dose children at 12 and 18 months after enrolment.

| Characteristic | 12 months after enrolment |  |  | 18 months after enrolment |  |  |
| --- | --- | --- | --- | --- | --- | --- |
|  | Fully immunised (n=452) | Incompletely immunised (n=2232) | p value | Fully immunised (n=1329) | Incompletely immunised (n=1355) | p value |
| Age at enrolment, months — median (IQR) | 21.4 (9.7–29.2) | 2.7 (1.9–4.4) | <0.001 | 3.1 (2.1–10.5) | 3 (2.0–6.7) | 0.008 |
| ≥1 year of age — n (%)<sup>†</sup> | 308 (68.1%) | 238 (10.7%) | <0.001 | 315 (23.7%) | 231 (17.0%) | <0.001 |
| Female — n (%) | 200 (44.2%) | 1076 (48.2%) | 0.134 | 608 (45.7%) | 668 (49.3%) | 0.069 |
| Residence: IDP — n (%) | 169 (37.4%) | 996 (44.6%) | 0.005 | 586 (44.1%) | 579 (42.7%) | 0.483 |
| Accelerated vaccination schedule — n (%) | 137 (30.3%) | 445 (19.9%) | <0.001 | 327 (24.6%) | 255 (18.8%) | <0.001 |
| Visits needed to become fully immunised — median (IQR)* | 3 (3–4) | 6 (5–6) | <0.001 | 6 (4–6) | 6 (5–6) | <0.001 |

\*Number of visits required based on age at enrolment and vaccination at a site that continuously used accelerated schedule or switched to routine schedule.
†Derived from age at enrolment
Characteristics of zero-dose children in the analytic cohort (≥18 months since enrolment) are compared by immunisation status at 12 and 18 months after enrolment. P values are from Fisher's exact test for categorical variables and the Wilcoxon rank-sum test for continuous variables. IDP: internally displaced persons. IQR: interquartile range.

Failure to return by 3, 6, 9, and 12 months was strongly associated with incomplete immunisation at 18 months, with positive predictive value increasing from 82% at 3 months to 99% at 12 months, and specificity from 93% to nearly 100% (Supplementary table 5). Sensitivity declined over time (33% to 23%), indicating that many children who ultimately did not achieve full immunisation had not yet failed to return at earlier time points.

Multivariate analyses. Among ZD children, several factors were associated with FI (Table 3. Multivariable models of factors associated with full immunisation among ZD children at 12 and 18 months). In Model A (baseline characteristics), older age at enrolment was associated with FI at 12 months (OR 1.11 per month, 95% CI 1.10–1.12), but not at 18 months (OR 1.00, 95% CI 0.99–1.01). Male sex was modestly associated with FI at both time points, while residence was not associated with FI. In Model B (programme delivery), greater vaccination opportunity was associated with higher odds of FI at 12 months (OR 2.09 per 0.5 increase in opportunity index, 95% CI 1.80–2.43), with a smaller effect at 18 months (OR 1.12, 95% CI 1.02–1.23)(Figure 2. Predicted probability of full immunisation among ZD children by age at enrolment, vaccination opportunity, and schedule). Model-based predictions (Figure 2) show that greater vaccination opportunity and use of accelerated schedules were associated with higher probability of FI, particularly at younger ages, with differences between groups narrowing by 18 months. In Supplementary Model C, which included programme retention, sustained programme engagement was strongly associated with FI (Supplementary Table 6).

**Table 3.** Factors associated with full immunisation among zero-dose children at 12 and 18 months.

A. Baseline predictors of full immunisation at 12 and 18 months
| Variable | 12 months OR (95% CI) | 18 months OR (95% CI) |
| --- | --- | --- |
| Age at enrolment (months) | 1.11 (1.10–1.12) | 1.00 (0.99–1.01) |
| Male sex | 1.30 (1.01–1.68) | 1.17 (1.00–1.37) |
| Village residence | 0.98 (0.75–1.27) | 0.95 (0.80–1.11) |

**Model B. Programme opportunity model for full immunisation at 12 and 18 months**
| Variable | 12 months OR (95% CI) | 18 months OR (95% CI) |
| --- | --- | --- |
| Age at enrolment (months) | 1.01 (0.99–1.04) | 0.98 (0.96–1.00) |
| Male sex | 1.28 (0.99–1.66) | 1.16 (0.99–1.36) |
| Village residence | 0.93 (0.71–1.23) | 0.94 (0.80–1.11) |
| Accelerated schedule | 3.00 (1.11–8.13) | 1.43 (0.77–2.66) |
| Vaccination opportunity (per 0.5 increase)* | 2.09 (1.80–2.43) | 1.12 (1.02–1.23) |

\*Vaccination opportunity was measured using the vaccination opportunity index, defined as the ratio of vaccination sessions available to the number of visits required to complete the vaccination schedule; ORs are presented per 0.5-unit increase in the index.

**Figure 2.**
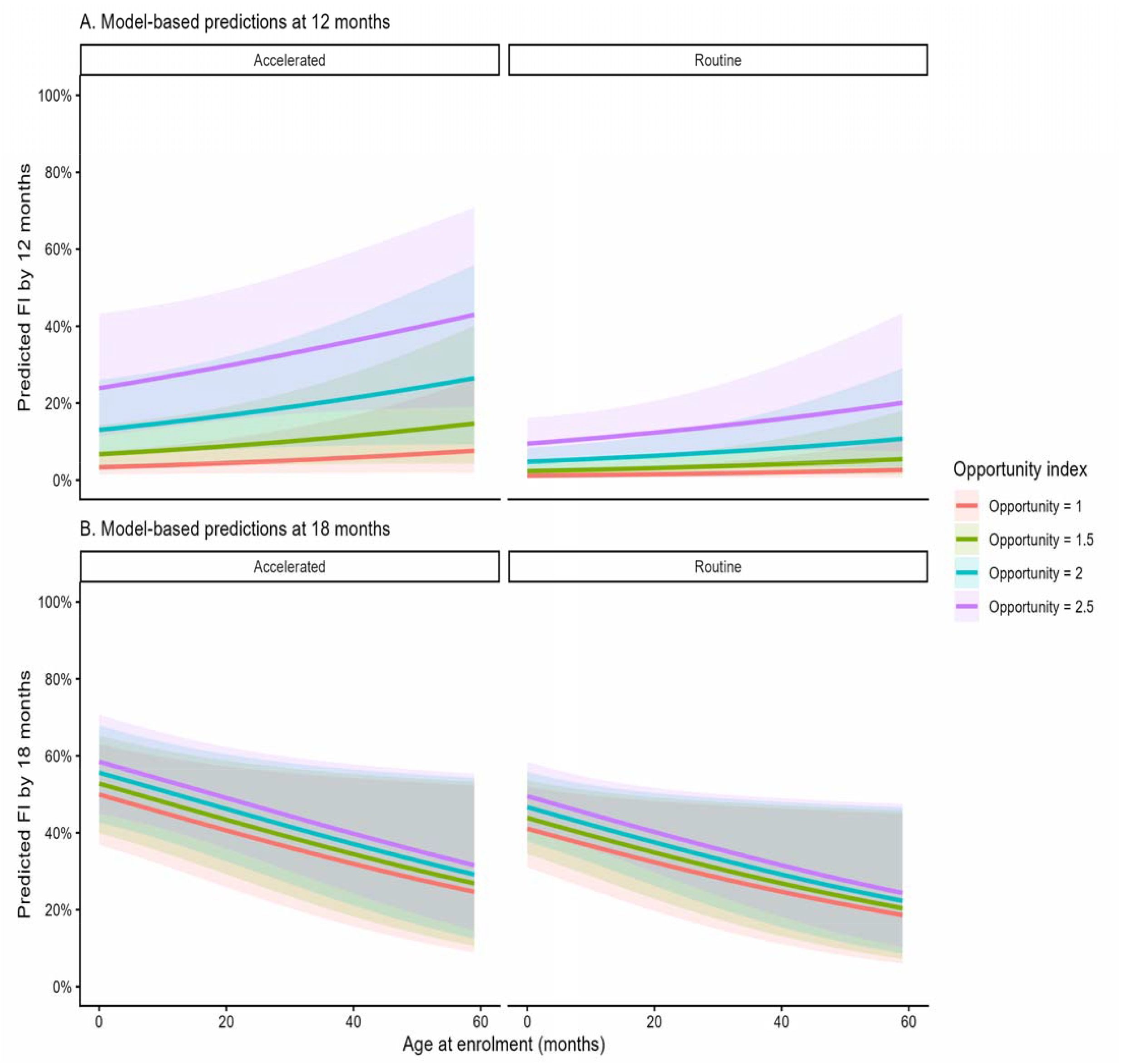
Predicted probability of full immunisation among zero-dose children by age at enrolment, vaccination opportunity, and schedule. Model-based predicted probabilities of achieving full immunisation (FI) are shown for zero-dose children across age at enrolment (months), stratified by vaccination opportunity and schedule. Predictions are derived from mixed-effects logistic regression models with a random intercept for site, holding sex and residence constant at their most common values. Panel A shows predicted probability of FI by 12 months after enrolment, and Panel B shows predicted probability of FI by 18 months. Lines represent levels of vaccination opportunity, defined as the number of vaccination sessions available relative to those required to complete the schedule. Shaded areas indicate 95% confidence intervals.

Cost analysis. Implementation was funded by the United Nations through intermediary non-governmental organizations. The cost per fully immunised ZD child was US$147 (Table 4. Comparative cost analysis of immunising children under programme vs Gavi/UNICEF program). Costs were driven primarily by vaccine prices (60%). Delivery costs—including staffing, transport, cold chain, supervision, and overhead—accounted for the remaining 40%. The largest delivery cost component was consultant costs, although most consultants worked pro bono. Sensitivity analysis using UNICEF/Gavi-equivalent prices reduced the hypothetical FI cost for the five vaccines to US$33. In sensitivity analysis, total programme expenditure would have fallen from US$901,184 to approximately US$155,000.

**Table 4.** Comparative cost analysis of immunising children under programme vs Gavi/UNICEF program Programme UNICEF*.

| Categories | <u>Programme</u> |  |  |  |  | <u>UNICEF*</u> |  |  |  |
| --- | --- | --- | --- | --- | --- | --- | --- | --- | --- |
|  | Per dose | Per fully immunised child | Per enrolled child | Total - all children | Percent | Per dose | Per fully immunised child | Total - all children | Percent |
| Vaccines | \$11.03 | \$110.30 | \$49.48 | \$542,046 | 60 | \$0.81 | \$8.10 | \$39,783 | 26 |
| Delivery costs<sup>†</sup> | \$4.92 | \$36.74 | \$22.05 | \$241,592 | 27 | \$2.34 | \$23.40 | \$114,997 | 74 |
| Cold chain | \$1.24 | \$12.42 | \$5.57 | \$61,042 | 7 | | | | |
| Vaccination session | \$0.81 | \$8.10 | \$3.63 | \$39,813 | 4 | | | | |
| Supervision | \$0.48 | \$4.79 | \$2.15 | \$23,550 | 3 | | | | |
| Consultants<sup>‡</sup> | \$2.38 | \$23.81 | \$10.68 | \$117,000 | 13 | | | | |
| Local capacity building | \$0.00 | \$0.04 | \$0.02 | \$188 | 0 | | | | |
| Overhead<sup>§</sup> | \$2.00 | \$20.04 | \$10.73 | \$117,546 | 13 | | | | |
| Total | \$15.95 | \$147.04 | | \$901,184 | 100 | \$3.15 | \$31.50 | \$154,780 | 100 |

\*Costs adapted from UNICEF<sup>31</sup>
† Delivery costs are the sum of the five categories below. For UNICEF, only the total available (individual categories not included)
‡ Consultants worked pro-bono. The value of their time was based on the amount paid to expatriate consultants by international non-governmental organizations in Myanmar.
§ Intermediary organization and Karenni Nurses Association.

Qualitative analyses. Findings from questionnaires and programme records indicated that community engagement and local coordination were central to service delivery. Community-based outreach, involvement of local leaders, and use of vaccination sites within villages and IDP camps increased awareness, trust, and demand for vaccination, and supported identification, referral, and return for subsequent doses. These mechanisms align with quantitative findings showing strong associations between vaccination opportunity, retention, and full immunisation (FI). As one nurse noted, “providing vaccinations increases community trust and allows people from remote and hard-to-reach areas to access immunisation more readily” (SWOT/respondent 2). High demand despite difficult access was evident, with one caregiver describing walking two days to reach a vaccination site.

Service delivery relied on coordination among community members, local authorities, and security actors, enabling vaccine transport and implementation in insecure settings. However, operational constraints were substantial. Transport challenges, particularly during the rainy season, and insecurity reduced the frequency of vaccination sessions and limited geographic coverage. As one report noted, “Because it is the rainy season… crossing the rivers could only be [done] by elephants… we had to reduce the vaccination sessions in Q3 and only 3 sites could get vaccinated” (quarterly report 2024/3).

Insecurity, population mobility, and limited access disrupted continuity of services, reducing both vaccination opportunities and sustained engagement. Dependence on external funding and intermediary organisations also constrained implementation, with procurement delays and vaccine shortages limiting availability, particularly for mobile or previously unreached populations. Together, these findings indicate that while community-based delivery enabled access in a conflict setting, service continuity remained highly sensitive to logistical, financial, and security constraints.

## DISCUSSION

Reconstructing routine immunisation in this active conflict setting was feasible but operationally challenging. Coverage fell short of the programme’s 12-month target (70%) among ZD children, but substantial gains were achieved over time. While 50% of ZD children were fully immunised by 18 months, the lower 12-month completion rate allowed identification of factors related to more rapid immunisation and highlighted a critical window for targeted follow-up and retention strategies. While causal inference is limited by the observational design, the findings are consistent with prior work and have implications for programme delivery in conflict-affected settings.

This study addresses several evidence gaps highlighted in reviews of the status of the Immunization Agenda 2030.^4 8^ First, it provides rare longitudinal data on ZD children in an active conflict setting, demonstrating both factors and obstacles to their vaccination. It documented immunisation how trajectories change over time, predictors of completion, the central role of retention, and costs.

Qualitative data showed how the quantitative data reflected underlying delivery processes—shaped by communityl71anchored, civill71societyl71led models that function amid state collapse—highlighting how contextl71specific operational realities in conflict settings align with IA2030’s emphasis on tailored strategies. The vaccination opportunity index provides a practical metric for measuring programme exposure in disrupted systems, contributing to improved monitoring approaches. Retention at 12 months may be a pragmatic indicator of programme engagement. Finally, the cost analysis adds to limited economic evidence on restoring routine immunisation in conflict-affected areas. Together, these findings strengthen the implementation evidence base needed to advance IA2030’s equity and ZD goals.

### Associations of FI

FI was higher among incompletely immunised children, reflecting fewer visits required for schedule completion. Among ZD children, vaccination opportunity and accelerated schedules influenced timing of completion, whereas the strong association of retention at 12 months likely reflects the dependence of FI on continued programme participation. Vaccination opportunity remained independently associated after adjustment, highlighting the importance of service availability.

The increasing positive predictive value of programme dropout over time indicates that children without return visits at later time points were increasingly likely to fail to achieve FI. Although many non-completers had not yet dropped out during earlier follow-up, later dropout was more strongly associated with non-completion, suggesting that programme dropout may serve as a useful operational indicator for identifying high-risk children and prioritizing retention efforts.

Vaccination opportunity and retention reflected delivery processes shaped by conflict, access, and governance. Community engagement was central to these dynamics. The previously reported^20^ strong demand for vaccination was central to the programme’s strengths. KNA’s longstanding partnerships with communities—including village authorities, religious leaders, and youth groups^20^—helped reduce access barriers and build trust, aligning with global recommendations that civil society organisations such as KNA contribute beyond mobilisation, operating across the immunisation delivery ecosystem, particularly where state authority is absent or contested.^2 4 32 33^

Return visits during the first year after enrolment provided a pragmatic indicator of programme continuity and were strongly associated with subsequent immunisation status. However, these measures provide only a partial assessment of programme performance, as they do not capture later dropout and may overestimate sustained coverage. In addition, some children classified as lost to follow-up may have continued vaccination outside the programme, leading to potential misclassification. Return visits should therefore be interpreted as operational indicators of continued programme engagement rather than proxies for full immunisation.

The premature transition from accelerated to routine schedules likely reflects changing conditions and programmatic decision-making. As security stabilised, some leaders judged routine schedules more appropriate for regular service delivery, reserving accelerated schedules for hard-to-reach or mobile populations. This aligns with our finding that accelerated delivery influenced the timing of completion rather than overall coverage.

Interpretation of FI should ideally distinguish between delivery failure and system-level constraints. While some children did not complete vaccination despite programme contact, reflecting access and delivery challenges, system constraints also played an important role. Periodic stockouts led to cancellation of vaccination sessions, directly reducing opportunities for vaccination and delaying completion. In addition, discontinuation of the Japanese encephalitis vaccine meant that full completion was not attainable for some. Together, these factors imply that observed coverage reflects both programme performance and structural limitations beyond programme control. External financing was essential to programme delivery. The cost per fully immunised ZD child (US$147) exceeded benchmarks from UNICEF and Gavi and was comparable to estimates from similar settings.^31 34^ Higher total costs were driven primarily by vaccine prices, reflecting limited access to pooled procurement and subsidised pricing. The use of single-dose and prefilled presentations further increased costs and cold chain requirements, placing additional strain on programme capacity.^35^ Delivery costs were broadly consistent with benchmarks once volunteer consultant contributions were excluded.^34^

### Implications for ZD strategies

Intermediary organisations provided essential funding but limited context-specific technical guidance, which may have contributed to unrealistic targets and operational inefficiencies. As reported elsewhere,^36^ intermediary organization’s inflexible policies and indicators designed without programme input often showed limited alignment with local implementation realities. In this context, standard EPI indicators based on vaccination by 12 months were insufficient, as they assume continuous service availability. Following the collapse of routine immunisation, extended age-based indicators were required to capture delayed access and programme recovery.

Expanding vaccination opportunities could further improve coverage, although this would increase demands on already stretched staff. This suggests dropout can serve as an early-warning indicator for targeted follow-up, with children who do not return within one year after enrolment actively traced and encouraged to re-engage, given the central role of retention.

These findings have implications for re-establishing routine immunisation in resistance-controlled areas of Myanmar. External funding will be required, as Gavi support is channelled through national governments, limiting access to subsidised vaccines for non-state providers.^17^ If experienced health workers with pre-existing immunisation skills are not present, capacity will need to be built through sustained investment in training and supportive supervision of community-based health workers, through strengthening local governance and coordination, and through ensuring reliable logistics and data systems to support safe, adaptive service delivery in insecure environments, as emphasised by World Health Organization guidance.^2 37 38^ Without such resources, our methods and results will be difficult to replicate.^17 19^

### Limitations

This evaluation has numerous limitations, many reflecting the challenges of conducting research on vaccination during conflict. First, the observational design limits causal inference, and associations may be affected by residual confounding, including age at enrolment and site-level operational factors. Population denominators were unavailable. Another limitation was our lack of information on intent to vaccinate (vaccine hesitancy, maternal education, decision making power for women), community access (travel expenses), and facility readiness.^39^ While greater facility readiness (e.g. integration of EPI with primary care) is recommended,^4^ this was not possible because the implementers did not have sufficient staff and donors did not prioritize integrated services. We did not study other demand-side factors such as religious beliefs, gender norms, lack of partner support, concerns about vaccine side effects, misconceptions, and caregiver knowledge of immunisation.^40^ Such data could not be collected because of the time and requirements for informed consent, which were deemed inappropriate in this low literacy setting.

To simplify analysis and increase statistical power, age criteria were not incorporated into definitions of ZD and incompletely immunised, although findings were robust in sensitivity analyses using standard definitions. Only five of the nine vaccines in Myanmar’s WHO EPI schedule were available, limiting comparability with routine programmes. Cost estimates, based on commercial vaccine pricing, are not generalisable to settings with access to global procurement mechanisms.

### Selection bias is likely

The vaccination status of children who never accessed the programme is unknown, limiting generalisability to the broader population of children. Some children classified as dropouts or incompletely immunised may have completed vaccination elsewhere. Use of a routine schedule at some sites was not random and was correlated with enrolment characteristics and site-level factors. Ethical constraints precluded randomised designs and limited collection of caregiver-level data (e.g., income, education, access barriers).

Termination of the JE vaccine, along with periodic vaccine stockouts and disruptions to service delivery, introduced measurement and implementation constraints that complicate interpretation of completion outcomes. In addition, time-to-event analyses were descriptive; more advanced approaches accounting for time-varying effects were not undertaken.

## Conclusion

In this conflict-affected setting, community-based delivery restored immunisation services among ZD children, fully immunising half within 18 months. The evaluation identified actionable steps to increase coverage: expanding vaccination opportunities, using accelerated schedules, and actively retaining children after enrolment. None of these is straightforward — implementing them requires transporting vaccines through active conflict zones, maintaining sessions despite insecurity, and tracing families across displacement camps. These demands underscore the need for sustained external financing and context-adapted support.41 42 Without durable peace and system rebuilding, accomplishments will remain fragile — but this programme demonstrates that the foundation can be laid even before those conditions exist.2 8 43

## Data Availability

All data produced in the present study are available upon reasonable request to the authors

## Competing interest

The first and last authors are independent researchers and, as consultants to Karenni Nurses Association, contributed to the design and implementation of the immunisation programme and this evaluation. These authors independently led data interpretation and manuscript preparation, for which they take full responsibility.

## Transparency Statement

the lead author (the manuscript’s guarantor) affirms that the manuscript is an honest, accurate, and transparent account of the study being reported; that no important aspects of the study have been omitted; and that any discrepancies from the study as originally planned (and, if relevant, registered) have been explained.

## Data sharing statement

The complete study database (without identifying information) and our R programme is available on request.

## Acknowledgments

We thank the families for the time, trust, and courage needed to bring their children to our vaccination sites. We also thank the Karenni Nurses Association (KNA) EPI and data management teams for their contributions throughout study implementation, data collection, analysis, and manuscript development. We are grateful to the community committees, youth groups, and local health workers who supported programme delivery and facilitated engagement with internally displaced and conflict-affected communities.

## Funding

Implementation was supported by United Nations through the Civil Health and Development Network (CHDN) through two agreements with the Karenni Nurses Association (KNA) for the project “Resources and Expertise to Advance Community Health in Karenni State.” These organizations had no role in study design, data interpretation, or writing of the manuscript. Some authors provided salary support for other authors to develop and collect data and review the manuscript.

## Notes

### Competing Interest Statement

The authors have declared no competing interest.

### Funding Statement

This study was funded by the United Nations

### Author Declarations

Ethics committee/IRB of Mae Tao Clinic gave ethical approval for this work

