## Supplementary for "Reconstruction of immunisation during conflict: A mixed-methods cohort evaluation of programme delivery and outcomes in Myanmar"

Supplementary Figures and Tables

The Karenni Nurses Association (KNA) was founded in 2022 by more than 160 Civil Disobedience Movement (CDM) nurses and midwives who left government service following the military coup and returned to Karenni State. Initially, many worked informally within their communities alongside other CDM health workers, but escalating conflict and successive waves of COVID-19 increased demand for health services. Early activities focused on providing care along the border and in displacement settings. With support from the Civil Health and Development Network, these efforts were later formalized into KNA to support internally displaced and conflict-affected populations.

One of the earliest identified needs was restoration of childhood vaccination services. KNA obtained vaccines and supplies through cross-border procurement from Thailand with support from international partners and transported them to displacement areas despite insecurity and access constraints. Drawing on prior experience in immunisation service delivery, KNA nurses and midwives established vaccination activities in multiple locations across Karenni State.

Over time, KNA expanded its activities alongside the broader disruption of Myanmar’s formal health system. Using the World Health Organization’s health system building blocks as an organizing framework, KNA developed teams for finance, procurement, logistics, service delivery, and workforce development. Activities included management of clinics, cold chain expansion, mobile outreach, emergency referral coordination, rehabilitation support, and delivery of primary care services to displaced populations.

To address workforce shortages, KNA also established Karenni Nursing University in 2022, offering a three-year Bachelor of Nursing Science programme through blended learning and cross-border partnerships. These activities were undertaken in collaboration with ethnic health organizations, civil society groups, and the National Unity Government, reflecting broader efforts to maintain essential health services in conflict-affected areas.

Supplementary Figure 1. Background of the Karenni Nurses Association

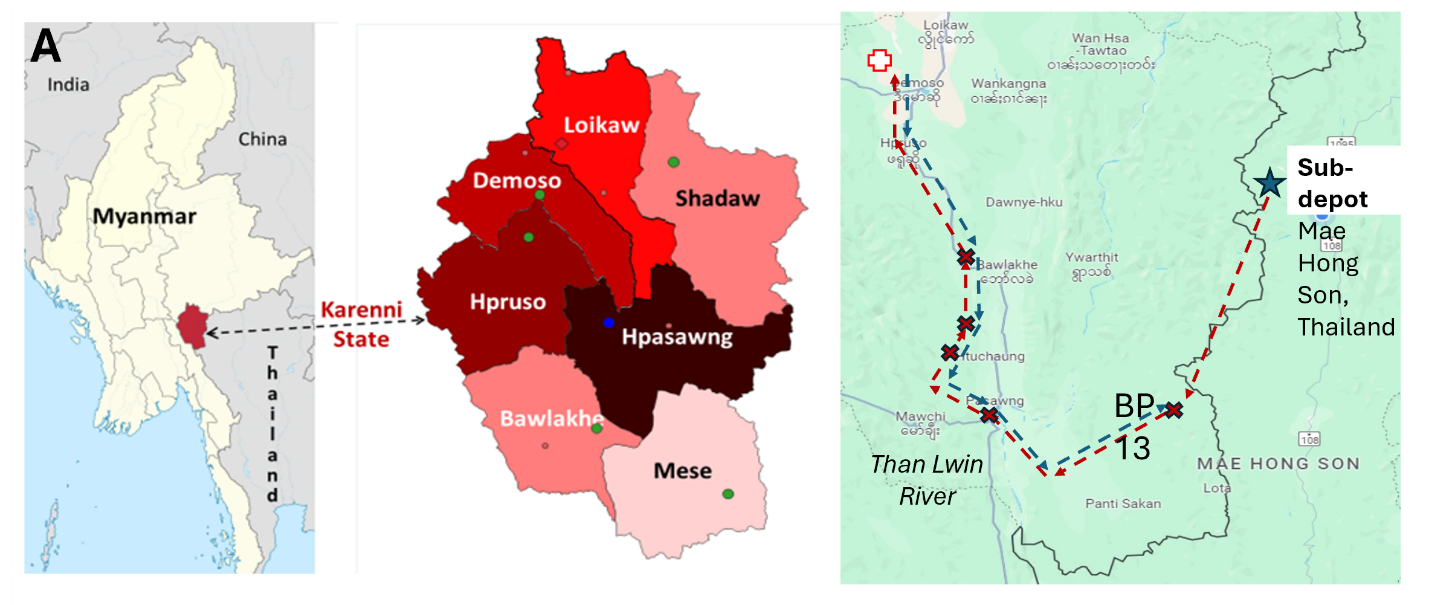

Supplementary Figure 2. Geographic setting and vaccine transport routes

A. Map showing Karenni State, township boundaries, immunisation sites, and the cross-border transport route from Thailand to the central cold chain facility. The figure illustrates geographic dispersion and access constraints in the study area.

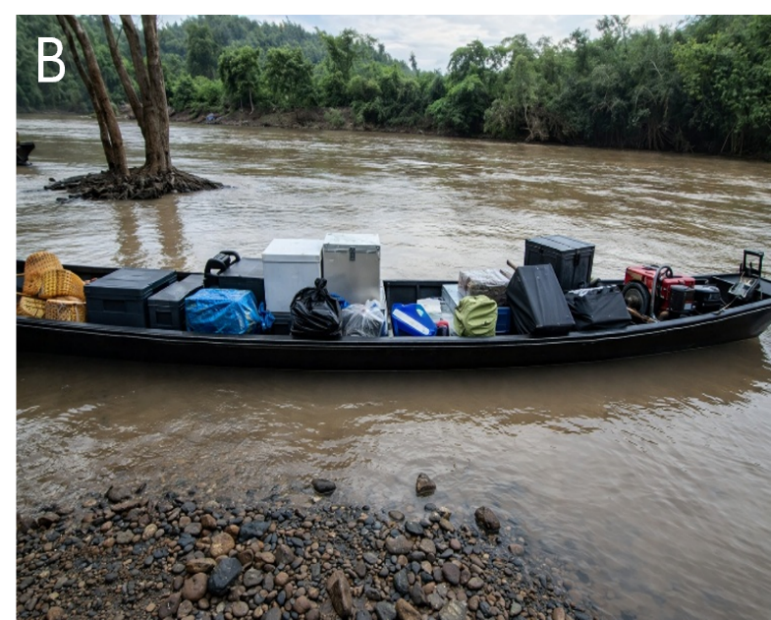

B. Photograph. Transport of vaccines (in white cold boxes) across a river by boat

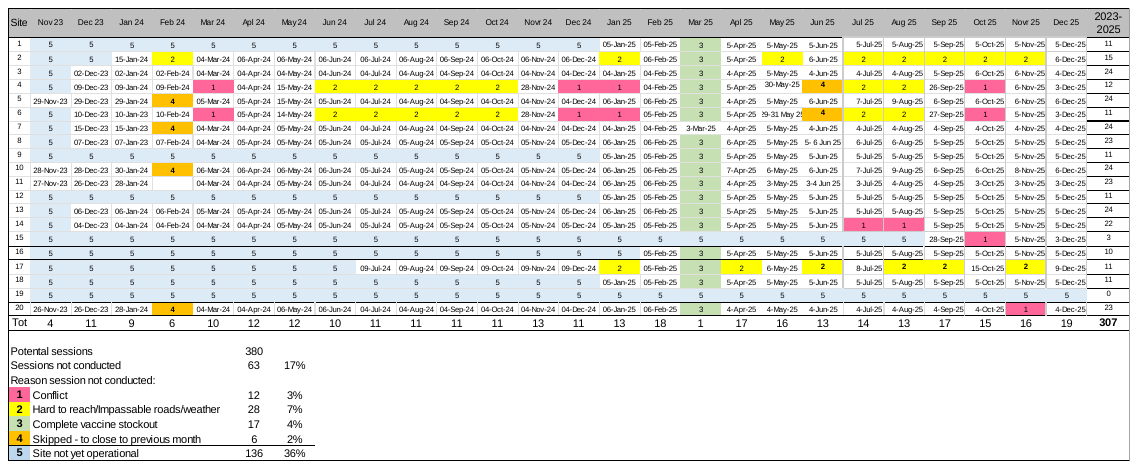

Supplementary Figure 3. Vaccination sites, dates of sessions, and the reason sessions were not conducted.

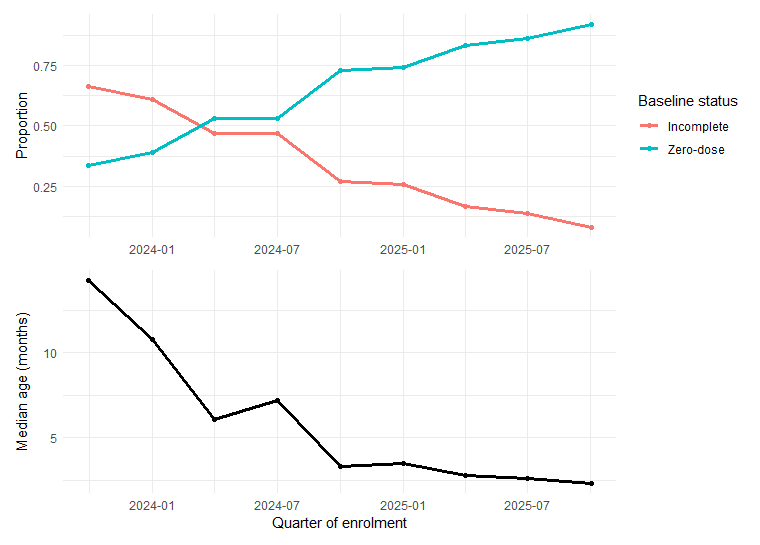

Supplementary Figure 4. Change in proportion of Zero-dose and median age over time.

Top panel shows the proportion of children enrolled each quarter who were zero-dose or incompletely immunised at enrolment. Bottom panel shows median age at enrolment. The increasing proportion of zero-dose children reflects progressively younger age at enrolment over time.

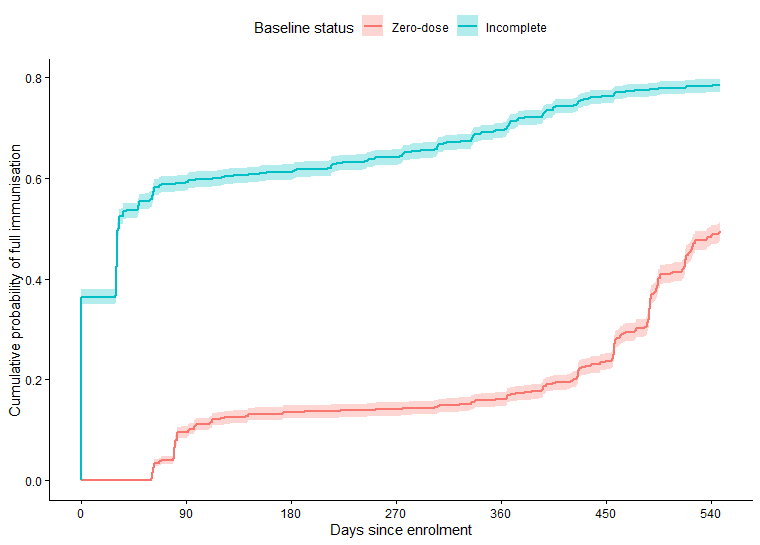

Supplementary Figure 5. Time from enrolment to full immunisation among zero-dose and incompletely immunised children.

Kaplan–Meier curves showing cumulative probability of full immunisation after enrolment among children eligible for 18-month follow-up. Full immunisation (FI) was defined based on completion of all required antigens using vaccination dates or documented prior doses; the date of full immunisation was taken as the date of the final dose administered within the programme. Curves are presented descriptively because zero-dose and incompletely immunised children differed at enrolment, including in the number of doses required to complete the schedule. Time to FI differed substantially by baseline status (log-rank p<0.001): children with incomplete immunisation at enrolment achieved FI rapidly (median 32 days, 95% CI 31–33), whereas the median was not reached among zero-dose children within 18 months. Differences between groups were most pronounced early and narrowed over time; the proportional hazards assumption was not met. Since the groups differ structurally at baseline, the KM comparison is descriptive of programme trajectories rather than a causal comparison of programme effect.

Supplementary table 1. Number of Visits Required for Full Immunization by Age at Enrolment

| Age at Enrolment | Accelerated Schedule | Routine Schedule |
| --- | --- | --- |
| <1.5 months | 6 | 6 |
| 1.5 to <7 months | 5 | 5 |
| 7 to <8 months | 4 | 5 |
| >= 8 to <9 months | 3 | 4 |
| >=9 months to <16 months | 3 | 4 |
| >=16 months | 3 | 3 |

Supplementary table 2. Indicators required by United Nations and reported at the end each quarter

| Indicator | Barrier to use |
| --- | --- |
| At least 1 dose of any vaccine on/before 5th birthday | No census available for denominators - Equivalent to total number of enrollees |
| Penta 1 before/on 5th birthday | No denominators available - Equivalent to total number of enrollees unless only BCG given |
| Penta 3 before/on 1st birthday | Insensitive to vaccination of older children enrolling |
| Penta 3 before/on 5th birthday* | No census available for denominators |
| MCV1 before/on 1st birthday | Insensitive to vaccination of older children enrolling |
| MCV1 before/on 5th birthday* | No census available for denominators |
| Penta 3, polio 3, MCV 2 before/on 5th birthday | JE encephalitis vaccine not included |
| Penta 1 to 3 dropout | Useful, but children may be vaccinated elsewhere |

*Proportion of population

Supplementary table 3. Baseline characteristics of children by eligibility for the analytic cohort

| **Characteristic** | **≥18 months since**  **enrolment (analytic cohort)** | **<18 months since**  **enrolment** | **p value** |
| --- | --- | --- | --- |
| N | 6563 | 6700 |  |
| Age at enrolment, median (IQR) | 10.5 (3.9–21.7) | 3.1 (2.2–10.8) | <0.001 |
| Age ≥1 year | 3052 (46.5%) | 1594 (23.8%) | <0.001 |
| Male | 3388 (51.6%) | 3475 (51.9%) | 0.773 |
| IDP | 3106 (47.3%) | 3276 (48.9%) | 0.070 |
| Zero-dose | 2684 (40.9%) | 4912 (73.3%) | <0.001 |

P values are from Fisher's exact test for categorical variables and the Wilcoxon rank-sum test for continuous variables.

Supplementary table 4. Baseline characteristics of children by immunisation status at enrolment and eligibility for the analytic cohort

|  | **Zero-dose** | | | **Incompletely immunised** | | |
| --- | --- | --- | --- | --- | --- | --- |
| **Characteristic** | **Enrolled for ≥18 months** | **Enrolled for <18 months** | **p** | **Enrolled for ≥18 months** | **Enrolled for <18 months** | **p** |
| N | 2684 | 4912 |  | 3879 | 1788 |  |
| Age at enrolment, months |  |  |  |  |  |  |
| Median (IQR) | 3.0 (2.1–8.4) | 2.6 (2.0–3.5) | <0.001 | 17.8 (9.5–26.2) | 18.9 (11.0–26.1) | <0.001 |
| ≥1 year — n (%) | 546 (20.3%) | 294 (6.0%) | <0.001 | 2506 (64.6%) | 1300 (72.7%) | <0.001 |
| Sex |  |  |  |  |  |  |
| Female — n (%) | 1276 (47.5%) | 2385 (48.6%) | 0.401 | 1899 (49.0%) | 839 (46.9%) | 0.161 |
| Male — n (%) | 1408 (52.5%) | 2527 (51.4%) |  | 1980 (51.0%) | 948 (53.0%) |  |
| Residence |  |  |  |  |  |  |
| IDP — n (%) | 1165 (43.4%) | 2347 (47.8%) | <0.001 | 1941 (50.0%) | 929 (52.0%) | 0.189 |
| Village — n (%) | 1519 (56.6%) | 2565 (52.2%) |  | 1938 (50.0%) | 859 (48.0%) |  |

IDP=Internally displaced person

P values are from Fisher's exact test for categorical variables and the Wilcoxon rank-sum test for continuous variables.

Supplementary Table 5. Return visit by time after enrolment as a predictor of immunisation status at 18 months among zero-dose children

| Time after enrolment | No return visit, n | Incom-pletely immunized, n | PPV, % | Return visits, n | Fully immunized, n | NPV, % | Specificity, % | Sensitivity, % |
| --- | --- | --- | --- | --- | --- | --- | --- | --- |
| 3 months | 542 | 444 | 81.9 | 2,104 | 1,231 | 58.5 | 92.6 | 33.7 |
| 6 months | 357 | 336 | 94.1 | 2,289 | 1,308 | 57.1 | 98.4 | 25.5 |
| 9 months | 315 | 307 | 97.5 | 2,331 | 1,321 | 56.7 | 99.4 | 23.3 |
| 12 months | 299 | 296 | 99.0 | 2,347 | 1,326 | 56.5 | 99.8 | 22.5 |

Legend. PPV = positive predictive value; NPV = negative predictive value. No return visit indicates children who had not attended any follow-up vaccination visit after programme initiation by the specified time point. Incompletely immunised at 18 months refers to children who had not achieved full immunisation by 18 months after enrolment. PPV is the proportion of children with no return visit who had not achieved full immunisation by 18 months. NPV is the proportion of children with at least one return visit who achieved full immunisation by 18 months. Specificity is the proportion of children who achieved full immunisation by 18 months who were correctly classified as having returned. Sensitivity is the proportion of children who were incompletely immunised at 18 months who were correctly classified as having no return visit.

Supplementary Table 6. Model C. Multivariable models of factors associated with full immunisation among zero-dose children at 18 months after enrolment

| **Variable** | **18 months OR (95% CI)** |
| --- | --- |
| Age at enrolment (months) | 0.99 (0.97–1.01) |
| Male sex | 1.12 (0.94–1.33) |
| Village residence | 0.93 (0.78–1.11) |
| Accelerated schedule | 1.48 (0.77–2.84) |
| Vaccination opportunities (per 0.5 increase) | 1.14 (1.03–1.27) |
| Retained at 12 months | 168.88 (53.64–531.65) |

Full immunisation was defined as receipt of all vaccines in the programme schedule.

The model included baseline demographic characteristics, programme delivery variables, and retention at 12 months, with study site included as a random intercept. Odds ratios (ORs) and 95% confidence intervals (CIs) are shown. Children retained at 12 months were defined as those returning for vaccination within 12 months of enrolment. Vaccination opportunity was measured using the vaccination opportunity index, defined as the ratio of vaccination sessions available to the number of visits required to complete the vaccination schedule; ORs are presented per 0.5-unit increase in the index.

The large odds ratio for retention at 12 months (OR 168.88, 95% CI 53.64–531.65) reflects quasi‑complete separation in the data: nearly all children who returned by 12 months subsequently achieved full immunisation at 18 months, whereas those who did not return rarely completed the schedule. This produces an inflated point estimate with wide uncertainty and should be interpreted as a strong indicator of programme continuity rather than a stable causal effect. The estimate primarily captures the structural dependence between early retention and later completion within the vaccination schedule.
